# High and increasing prevalence of SARS-CoV-2 swab positivity in England during end September beginning October 2020: REACT-1 round 5 updated report

**DOI:** 10.1101/2020.10.12.20211227

**Authors:** Steven Riley, Kylie E. C. Ainslie, Oliver Eales, Caroline E. Walters, Haowei Wang, Christina Atchison, Claudio Fronterre, Peter J. Diggle, Deborah Ashby, Christl A. Donnelly, Graham Cooke, Wendy Barclay, Helen Ward, Ara Darzi, Paul Elliott

## Abstract

**Background:** REACT-1 is quantifying prevalence of SARS-CoV-2 infection among random samples of the population in England based on PCR testing of self-administered nose and throat swabs. Here we report results from the fifth round of observations for swabs collected from the 18th September to 5th October 2020. This report updates and should be read alongside our round 5 interim report.

**Methods:** Representative samples of the population aged 5 years and over in England with sample size ranging from 120,000 to 175,000 people at each round. Prevalence of PCR-confirmed SARS-CoV-2 infection, estimation of reproduction number (R) and time trends between and within rounds using exponential growth or decay models.

**Results:** 175,000 volunteers tested across England between 18th September and 5th October. Findings show a national prevalence of 0.60% (95% confidence interval 0.55%, 0.71%) and doubling of the virus every 29 (17, 84) days in England corresponding to an estimated national R of 1.16 (1.05, 1.27). These results correspond to 1 in 170 people currently swab-positive for the virus and approximately 45,000 new infections each day. At regional level, the highest prevalence is in the North West, Yorkshire and The Humber and the North East with strongest regional growth in North West, Yorkshire and The Humber and West Midlands.

**Conclusion:** Rapid growth has led to high prevalence of SARS-CoV-2 virus in England, with highest rates in the North of England. Prevalence has increased in all age groups, including those at highest risk. Improved compliance with existing policy and, as necessary, additional interventions are required to control the spread of SARS-CoV-2 in the community and limit the numbers of hospital admissions and deaths from COVID-19.

## Introduction

The second wave of the COVID-19 pandemic in England continues to cause increased hospitalizations, ICU bed admissions and deaths [1]. The REal-time Assessment of Community Transmission-1 (REACT-1) study has been quantifying the community prevalence of SARS-CoV-2 virus over repeated rounds of data collection among separate samples of 120,000 to 175,000 people since May 2020 [2,3]. We previously reported a rapid increase in infections in England between round 4 (mid August to early September 2020) [4] and interim results for round 5 (18th to 26th September 2020) [5]. Here, we report results for the full period of data collection in round 5, to 5th October 2020.

## Methods

Methods of REACT-1 have been described previously [2,3]. Briefly, data collection is based on repeated random samples of the population at ages five years and over. Sampling uses the National Health Service (NHS) list of patients registered with a general practitioner, stratified by the 315 lower-tier local authorities in England. Participants provide a self-administered nose and throat swab (parent / guardian administered for children ages five to 12 years) and complete an online or telephone questionnaire. Swabs are sent on a cold chain to the laboratory for PCR testing. We estimated prevalence by dividing numbers of swab-positive results by numbers tested, and at national level reweighted prevalence estimates to be representative of the population of England. We analysed time trends between and within rounds using exponential growth or decay models, assuming numbers of positive swabs arose from a binomial distribution. We estimated the reproduction number R and doubling times, assuming the generation time followed a gamma distribution (mean 6.29 days, standard deviation of 4.2) [6]; R was estimated both between rounds and separately within each round of data collection. We used multivariable logistic regression to obtain adjusted odds ratios and 95% confidence intervals for associations of swab positivity with covariates.

Research ethics approval was obtained from the South Central-Berkshire B Research Ethics Committee (IRAS ID: 283787).

## Results

These results for round 5 update those presented in our interim analysis but with increased precision reflecting the larger sample size [5]. From 174,949 swab results we found 824 positive samples for a weighted national average prevalence of 0.60% (95% confidence interval 0.55%, 0.71%) (Table 1, Figure 1).

**Table 1.** Prevalence of swab-positivity across all five rounds of REACT-1.

| Round | Tested swabs | Positive swabs | Unweighted prevalence (95% CI) | Weighted prevalence (95% CI) | First sample | Last sample |
| --- | --- | --- | --- | --- | --- | --- |
| 1 | 120,620 | 159 | 0.13% (0.11%, 0.15%) | 0.16% (0.13%, 0.19%) | 1/5/2020 | 1/6/2020 |
| 2 | 159,199 | 123 | 0.077% (0.065%, 0.092%) | 0.088% (0.068%, 0.11%) | 19/6/2020 | 7/7/2020 |
| 3 | 161,560 | 54 | 0.033% (0.025%, 0.043%) | 0.040% (0.027%, 0.053%) | 24/7/2020 | 11/8/2020 |
| 4 | 154,325 | 137 | 0.089% (0.075%, 0.11%) | 0.13% (0.096%, 0.15%) | 20/8/2020 | 8/9/2020 |
| 5 | 174,949 | 824 | 0.51% (0.46%, 0.56%) | 0.60% (0.55%, 0.71%) | 18/9/2020 | 5/10/2020 |

**Figure 1.**
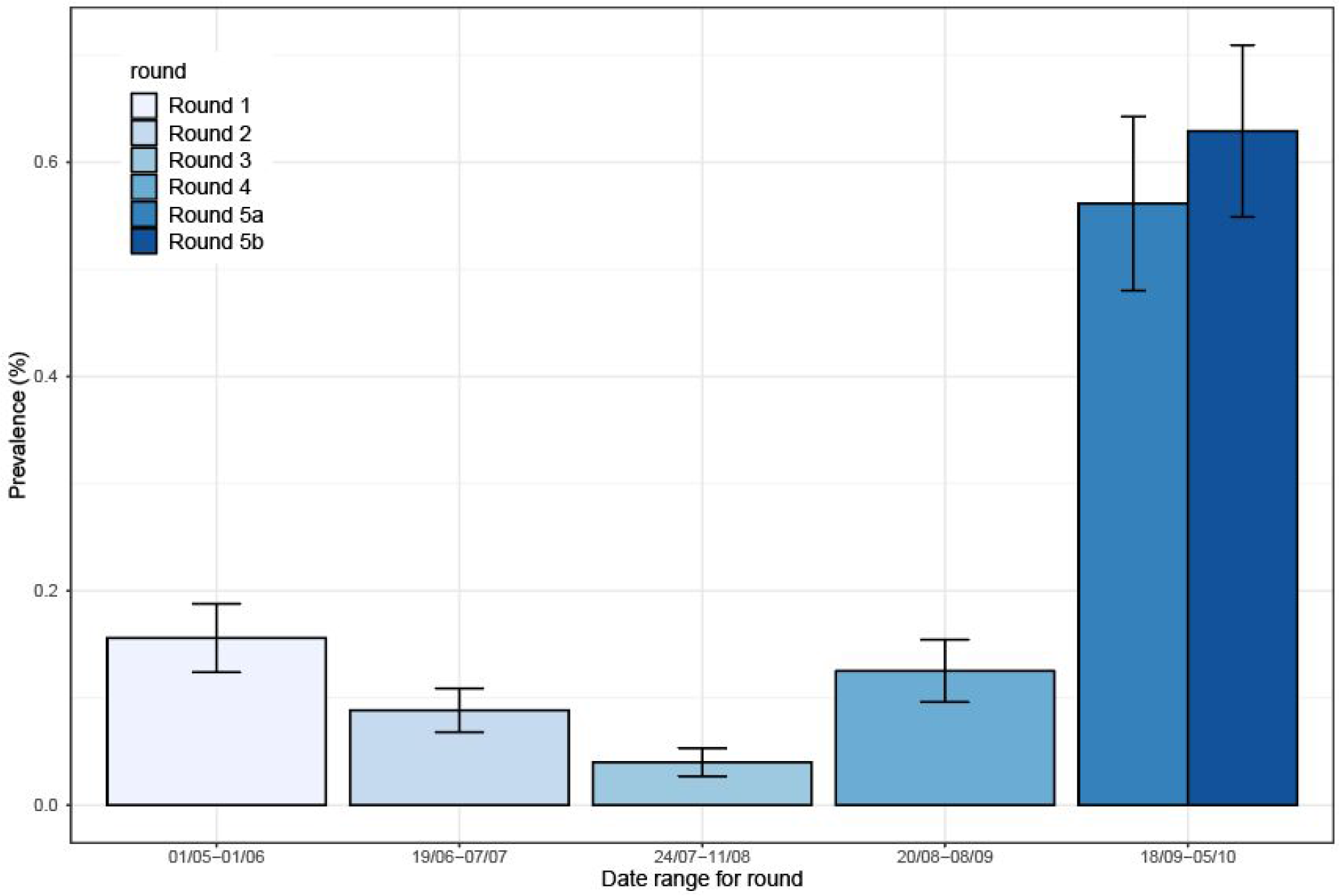
Weighted prevalence for 4 complete rounds of REACT-1 and round 5 split into 2 halves. Results from the first half of round 5 were reported in Ref [5].

Overall, using data only from round 5, at the national level, we observe a growing epidemic with a doubling time of 29 (17, 84) days and a corresponding R number of 1.16 (1.05,1.27) (Table 2, Figure 2). This represents a modest deceleration from the doubling time of 13 (11,14) days estimated using data from both rounds 4 and 5, which corresponds to an R value of 1.39 (1.34, 1.43).

**Table 2.** National estimates of growth rate, doubling time and reproduction number for round 5 and for rounds 4 and 5 together.

| Round | Round | Growth rate ($r$, 1/days) | Reproduction number | $P(R>1)$ | Doubling time (+) /<br>halving time (-) (days) |
| --- | --- | --- | --- | --- | --- |
| All positives | 4 and 5 | 0.055 ( 0.050 , 0.061 ) | 1.39 ( 1.34 , 1.43 ) | 1.00 | 13 ( 14 , 11 ) |
|  | 5 | 0.024 ( 0.008 , 0.040 ) | 1.16 ( 1.05 , 1.27 ) | 1.00 | 29 ( 84 , 17 ) |
| Asymptomatic positives | 4 and 5 | 0.045 ( 0.037 , 0.053 ) | 1.31 ( 1.25 , 1.37 ) | 1.00 | 15 ( 19 , 13 ) |
|  | 5 | 0.005 ( -0.021 , 0.029 ) | 1.03 ( 0.87 , 1.19 ) | 0.65 | 145 ( -34 , 24 ) |
| Positive for both gene<br>targets | 4 and 5 | 0.060 ( 0.054 , 0.067 ) | 1.42 ( 1.37 , 1.48 ) | 1.00 | 12 ( 13 , 10 ) |
|  | 5 | 0.019 ( 0.001 , 0.037 ) | 1.12 ( 1.00 , 1.25 ) | 0.98 | 37 ( 1010 , 19 ) |
| Positive for both gene<br>targets or N gene CT < 35* | 4 and 5 | 0.056 ( 0.050 , 0.063 ) | 1.40 ( 1.35 , 1.44 ) | 1.00 | 12 ( 14 , 11 ) |
|  | 5 | 0.023 ( 0.006 , 0.040 ) | 1.15 ( 1.04 , 1.27 ) | 1.00 | 30 ( 120 , 17 ) |

\* All positives use a CT threshold of 37

**Figure 2.**
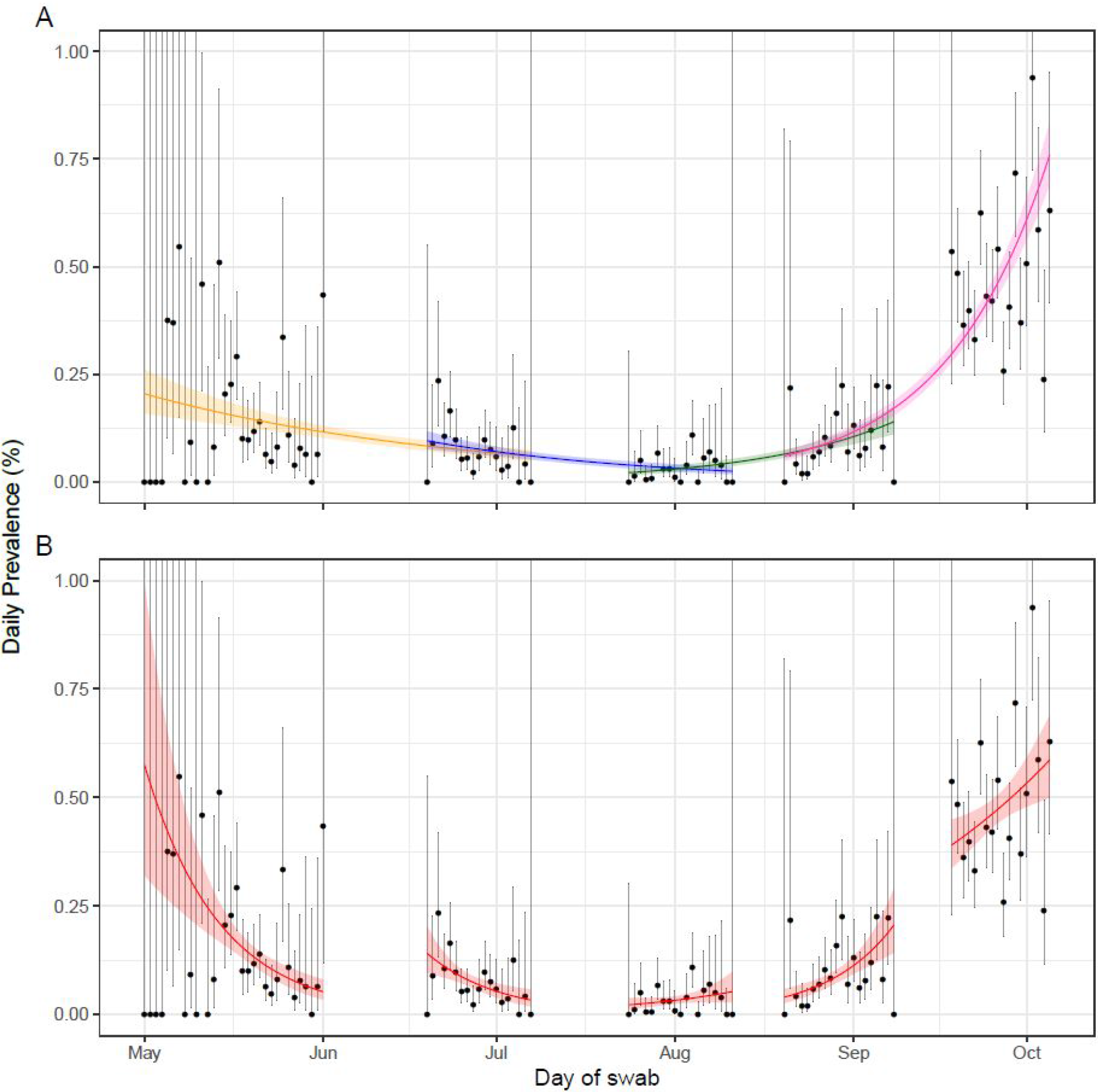
Constant growth rate models fit to REACT-1 data for sequential and individual rounds. **A** models fit to REACT-1 data for sequential rounds; 1 and 2 (yellow), 2 and 3 (blue), 3 and 4 (green) and 4 and 5 (pink). **B** models fit to individual rounds only (red). Vertical lines show 95% confidence intervals for observed prevalence (black points). Shaded regions show 95% posterior credible intervals for growth models. Underlying data are provided in the [supporting data](https://docs.google.com/spreadsheets/d/1qtbl4LTtCKZNDpDRuHAxNFeJIlYi1SeH_vvzfZ5sm_k/edit?usp=sharing).

We found marked heterogeneity in prevalence in round 5 at regional level (Table 3, Figure 3). Unweighted prevalence was highest in: North West, 1.02% (0.89%, 1.17%); North East, 0.91% (0.71%, 1.17%) and Yorkshire and The Humber 0.63% (0.51%, 0.80%). Unweighted prevalence was lowest in the South East at 0.26% (0.22%, 0.32%). London had an intermediate prevalence of 0.45% (0.36%, 0.56%).

**Table 3.**
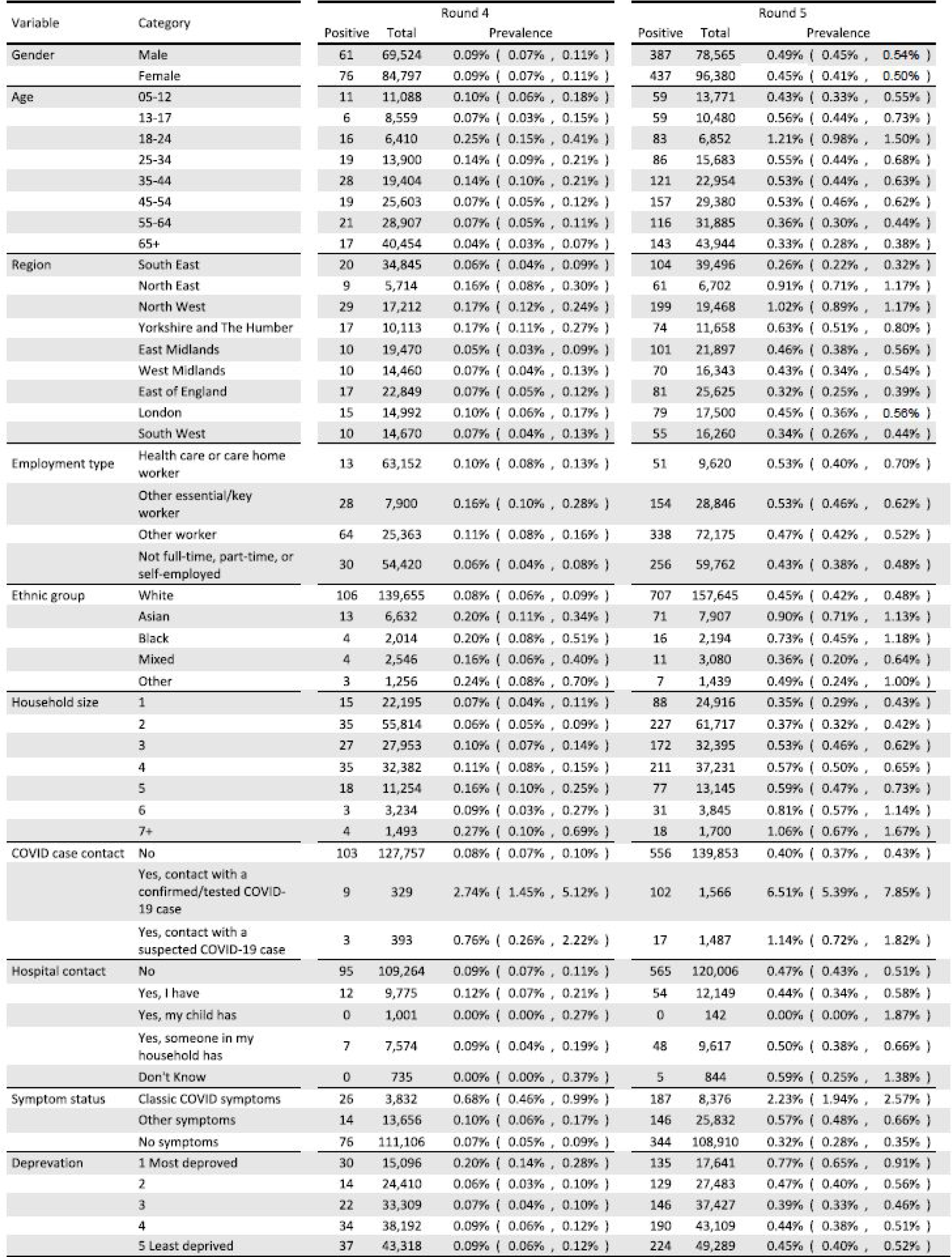
Unweighted prevalence of swab-positivity by variable and category for rounds 4 and 5 of REACT-1.

**Figure 3.**
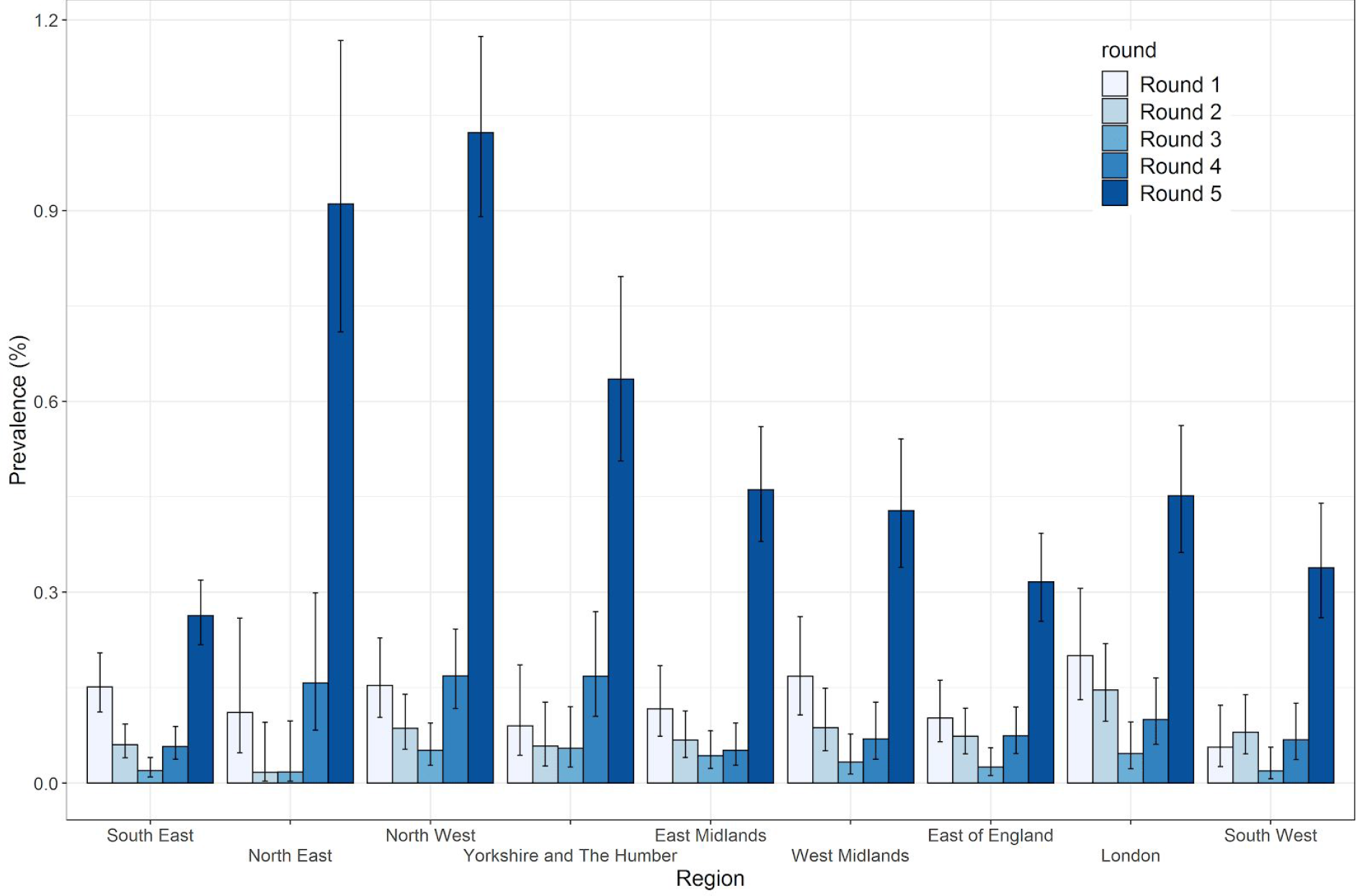
Unweighted prevalence and 95% confidence intervals of swab-positivity by region by round.

Using data only from round 5, we also found marked heterogeneity in growth patterns and R values at regional level (Table 4, Figure 4). We found greater than 95% probability that regional epidemics were growing in: North West, R = 1.27 (1.04, 1.53); Yorkshire and the Humber, R = 1.37 (1.00, 1.80); and West Midlands, R = 1.33 (0.97, 1.78). For London we estimated an R value of 0.97 (0.68, 1.31).

**Table 4.** Regional estimates of growth rate and reproduction number for round 5.

| Region | Growth rate (r) | Reproduction number | P(R>1) |
| --- | --- | --- | --- |
| East Midlands | 0.030 ( -0.016 , 0.074 ) | 1.20 ( 0.90 , 1.54 ) | 0.90 |
| West Midlands | 0.048 ( -0.004 , 0.103 ) | 1.33 ( 0.97 , 1.78 ) | 0.96 |
| East of England | 0.007 ( -0.045 , 0.058 ) | 1.05 ( 0.73 , 1.41 ) | 0.61 |
| London | -0.004 ( -0.056 , 0.046 ) | 0.97 ( 0.68 , 1.31 ) | 0.44 |
| North West | 0.040 ( 0.006 , 0.073 ) | 1.27 ( 1.04 , 1.53 ) | 0.99 |
| North East | 0.008 ( -0.051 , 0.066 ) | 1.05 ( 0.70 , 1.47 ) | 0.60 |
| South East | 0.033 ( -0.014 , 0.078 ) | 1.22 ( 0.91 , 1.57 ) | 0.92 |
| South West | 0.025 ( -0.034 , 0.079 ) | 1.16 ( 0.80 , 1.58 ) | 0.79 |
| Yorkshire and<br>The Humber | 0.053 ( -0.001 , 0.106 ) | 1.37 ( 1.00 , 1.80 ) | 0.97 |

**Figure 4.**
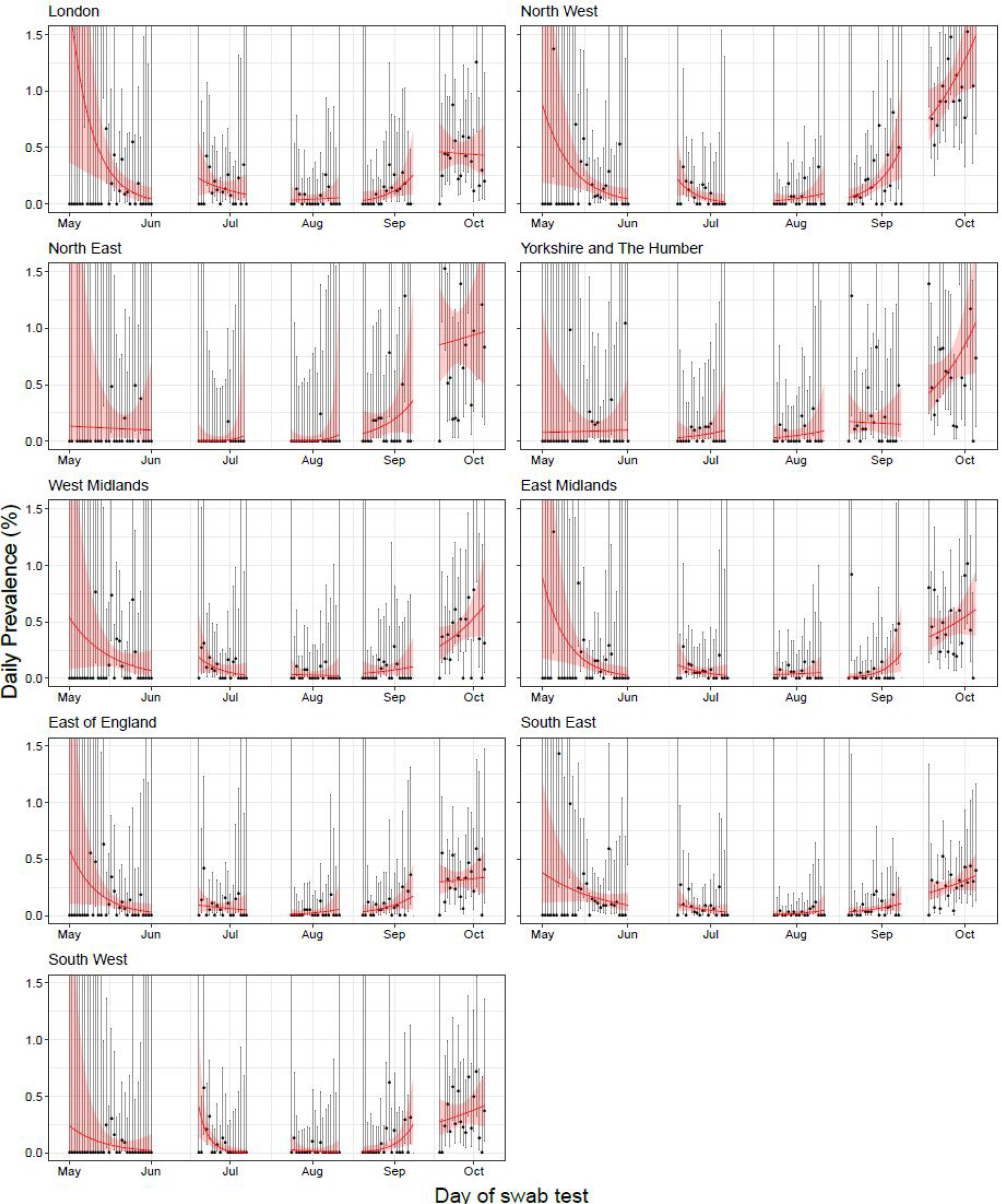
Constant growth rate models fit to REACT-1 data by region for individual rounds only. Vertical lines show 95% confidence intervals for observed prevalence (black points). Shaded regions show 95% posterior credible intervals for growth models. Underlying data are provided in the [supporting data](https://docs.google.com/spreadsheets/d/1qtbl4LTtCKZNDpDRuHAxNFeJIlYi1SeH_vvzfZ5sm_k/edit?usp=sharing).

The large number of positive swabs in this round permit an informative map of observed prevalence by lower tier local authority (LTLA, Figure 5). As expected from regional prevalence, the highest prevalence local authorities were mainly in the North and Midlands. The supporting data for this map are available by LTLA in the [supporting data](https://docs.google.com/spreadsheets/d/1qtbl4LTtCKZNDpDRuHAxNFeJIlYi1SeH_vvzfZ5sm_k/edit?usp=sharing).

**Figure 5.**
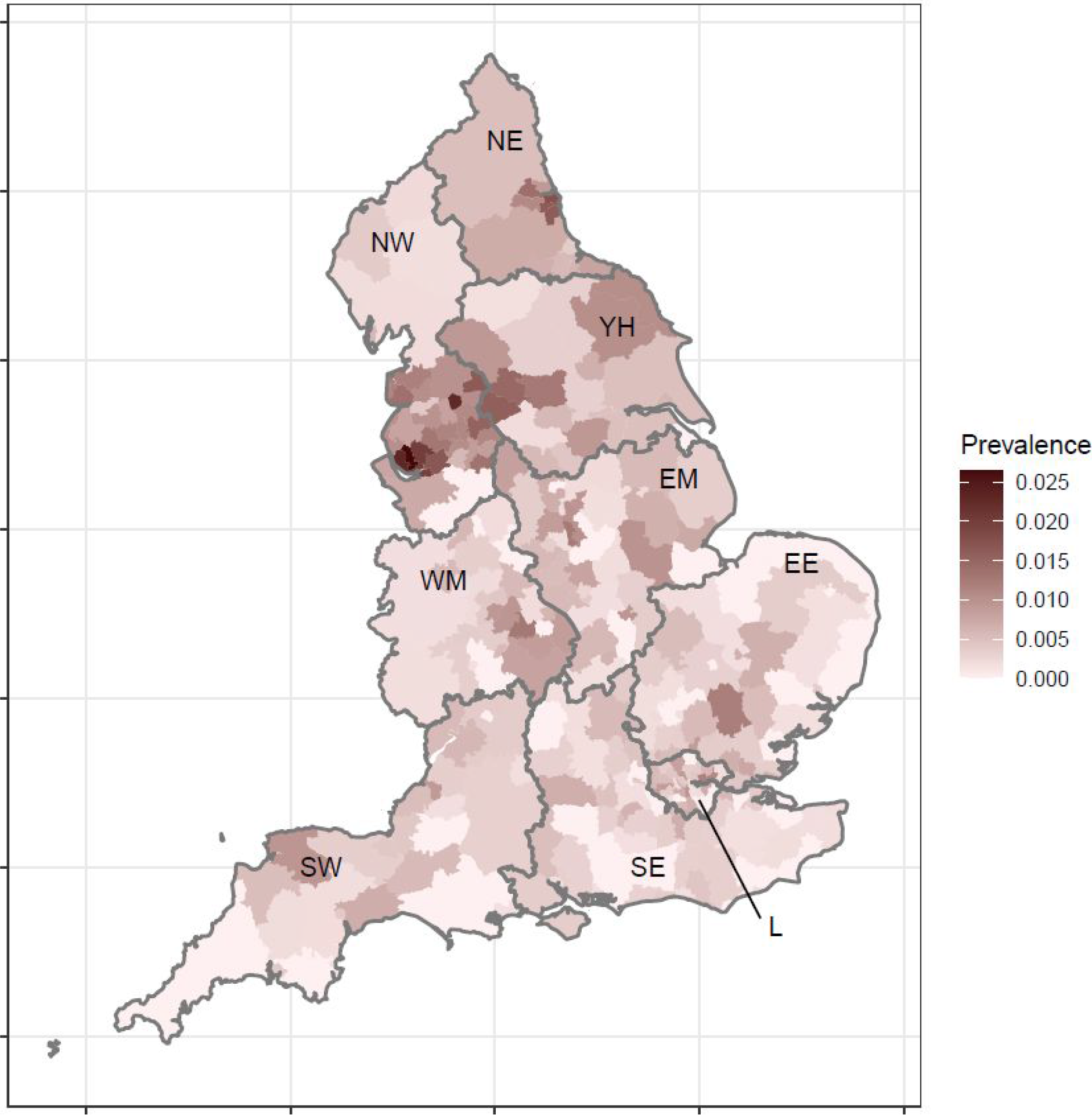
Prevalence of swab-positivity by lower-tier local authority (LTLA). NE = North East, NW = North West, YH = Yorkshire and The Humber, EM = East Midlands, WM = West Midlands, EE = East of England, L = London, SE = South East, SW = South West. Note that these individual LTLA estimates are based on small numbers. Underlying data and confidence intervals are provided in the [supporting data](https://docs.google.com/spreadsheets/d/1qtbl4LTtCKZNDpDRuHAxNFeJIlYi1SeH_vvzfZ5sm_k/edit?usp=sharing).

Since the previous round, unweighted prevalence increased in all age groups (Table 3, Figure 6). The highest prevalence was amongst the 18-to 24-year olds at 1.21% (0.98%, 1.50%). For those ages 65 and over it increased 8-fold between the previous and present round from 0.04% (0.03%, 0.07%) to 0.33% (0.28%, 0.38%).

**Figure 6.**
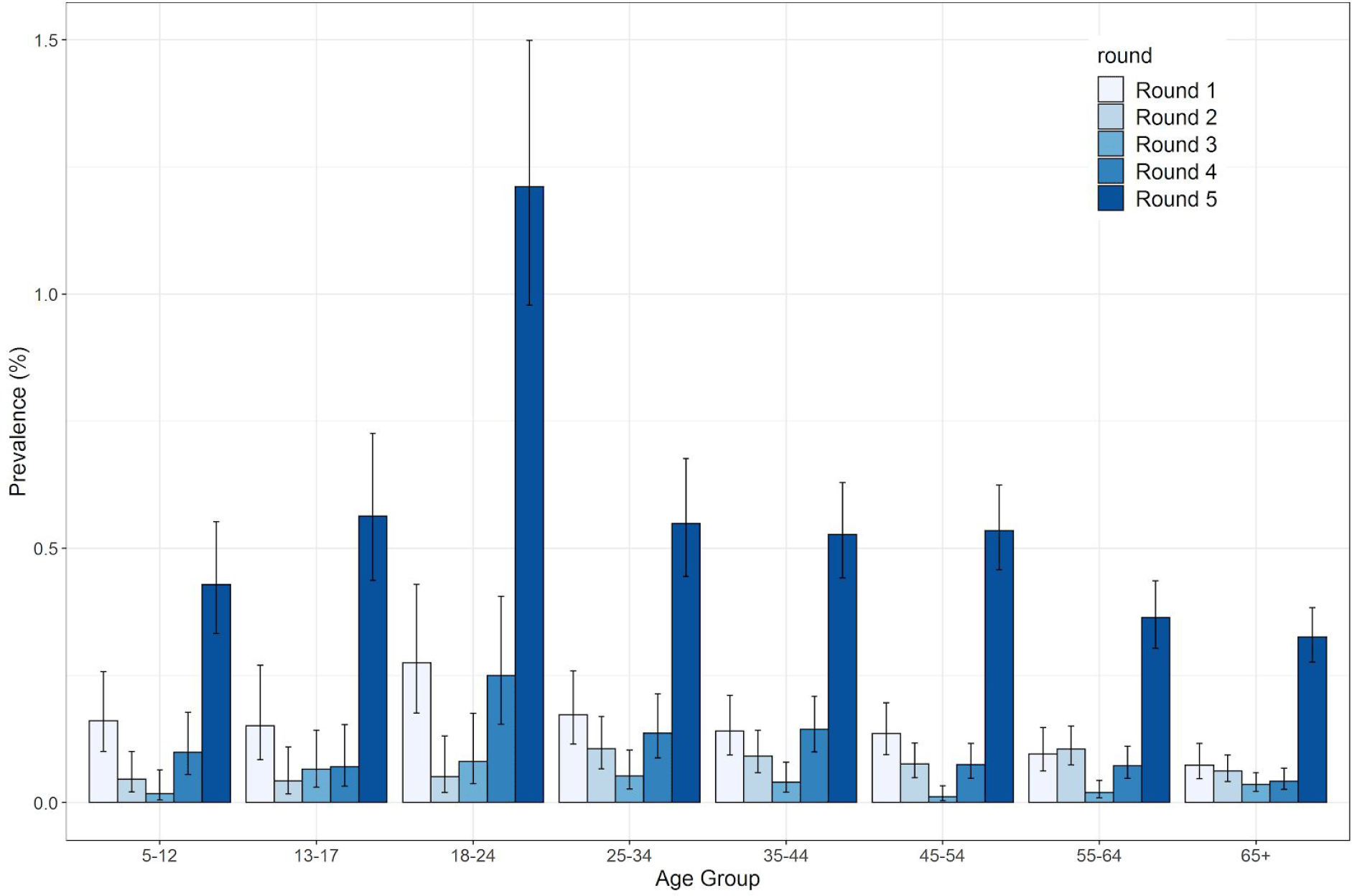
Unweighted prevalence and 95% confidence intervals of swab-positivity by age by round.

Unweighted prevalence was highest in participants of Asian and Black ethnicity at 0.90% (0.71%, 1.13%) and 0.73% (0.45%, 1.18%) respectively (Table 3); this compares with 0.45% (0.42%, 0.48%) in white participants. Unweighted prevalence was highest in those living in large households. For those living in households of size 6 or 7+ prevalence was 0.81% (0.57%, 1.14%) and 1.06% (0.67%, 1.67%) respectively (Table 3); this compares with 0.35% (0.29%, 0.43%) in single occupancy households. Unweighted prevalence was highest in participants who lived in the most deprived areas at 0.77% (0.65%, 0.91%) (Table 3)

We used logistic regression to estimate jointly-adjusted odds ratios for: age, region, ethnicity, work type, household size and quintile of deprivation (Table 5, Figure 7). The effect of deprivation was substantially reduced in the jointly adjusted model.

**Table 5.** Odds ratios for logistic regression models of swab-positivity for REACT-1 round 5.

| Variable | Category | Unadjusted | Adjusted only for age and sex | Jointly adjusted |
| --- | --- | --- | --- | --- |
| Gender | Male | Ref | Ref | Ref |
|  | Female | 0.92 [0.80,1.06] | 0.90 [0.78,1.03] | 0.90 [0.78,1.04] |
| Age Group | 5-12 | 0.81 [0.59,1.11] | 0.80 [0.59,1.10] | 0.71 [0.51,0.99] |
|  | 13-17 | 1.07 [0.78,1.46] | 1.06 [0.77,1.45] | 1.02 [0.70,1.47] |
|  | 18-24 | 2.31 [1.75,3.06] | 2.31 [1.75,3.06] | 2.17 [1.61,2.92] |
|  | 25-34 | 1.04 [0.79,1.37] | 1.04 [0.79,1.38] | 1.08 [0.81,1.44] |
|  | 35-44 | Ref | Ref | Ref |
|  | 45-54 | 1.01 [0.80,1.29] | 1.01 [0.80,1.28] | 1.05 [0.82,1.35] |
|  | 55-64 | 0.69 [0.53,0.89] | 0.69 [0.53,0.88] | 0.76 [0.58,1.00] |
|  | 65+ | 0.62 [0.48,0.79] | 0.61 [0.48,0.78] | 0.72 [0.53,0.97] |
| Region | North East | 3.48 [2.53,4.78] | 3.46 [2.52,4.76] | 3.63 [2.61,5.07] |
|  | North West | 3.91 [3.08,4.96] | 3.92 [3.09,4.98] | 4.01 [3.12,5.16] |
|  | Yorkshire and The Humber | 2.42 [1.79,3.26] | 2.44 [1.81,3.29] | 2.53 [1.85,3.46] |
|  | East Midlands | 1.76 [1.33,2.31] | 1.76 [1.34,2.32] | 1.83 [1.38,2.43] |
|  | West Midlands | 1.63 [1.20,2.21] | 1.62 [1.20,2.20] | 1.69 [1.23,2.31] |
|  | East of England | 1.20 [0.90,1.61] | 1.19 [0.89,1.60] | 1.20 [0.88,1.62] |
|  | London | 1.72 [1.28,2.30] | 1.63 [1.21,2.18] | 1.46 [1.06,2.00] |
|  | South East | Ref | Ref | Ref |
|  | South West | 1.29 [0.93,1.78] | 1.32 [0.95,1.83] | 1.35 [0.95,1.91] |
| Key Worker Status | HCW/CHW | 1.23 [0.90,1.66] | 1.23 [0.90,1.67] | 1.07 [0.78,1.46] |
|  | Key worker (other) - public facing | 1.21 [0.96,1.53] | 1.15 [0.91,1.45] | 1.08 [0.85,1.36] |
|  | Key worker (other) - not public facing | 1.25 [0.92,1.71] | 1.17 [0.85,1.60] | 1.06 [0.78,1.45] |
|  | Other worker - public facing | 1.33 [1.03,1.71] | 1.28 [0.99,1.65] | 1.22 [0.94,1.57] |
|  | Other worker - not public facing | Ref | Ref | Ref |
|  | Not FT, PT, SE | 0.99 [0.83,1.18] | 1.16 [0.94,1.41] | 1.07 [0.87,1.31] |
| Ethnicity | Asian | 2.01 [1.57,2.57] | 1.81 [1.41,2.32] | 1.92 [1.47,2.52] |
|  | Black | 1.63 [0.99,2.68] | 1.51 [0.92,2.49] | 1.64 [0.97,2.79] |
|  | Mixed | 0.80 [0.44,1.44] | 0.70 [0.39,1.28] | 0.80 [0.42,1.49] |
|  | Other | 1.09 [0.51,2.29] | 1.03 [0.49,2.17] | 1.19 [0.56,2.54] |
|  | White | Ref | Ref | Ref |
| Household Size | 1-2 | Ref | Ref | Ref |
|  | 2-5 | 1.53 [1.33,1.77] | 1.28 [1.08,1.53] | 1.24 [1.04,1.49] |
|  | 6+ | 2.44 [1.81,3.31] | 2.01 [1.46,2.78] | 1.89 [1.34,2.65] |
| Deprivation Index Quintile | 1 | Ref | Ref | Ref |
|  | 2 | 0.61 [0.48,0.78] | 0.63 [0.49,0.80] | 0.81 [0.63,1.05] |
|  | 3 | 0.51 [0.40,0.64] | 0.53 [0.42,0.67] | 0.79 [0.62,1.02] |
|  | 4 | 0.57 [0.46,0.72] | 0.61 [0.49,0.76] | 0.89 [0.70,1.13] |
|  | 5 | 0.59 [0.48,0.73] | 0.63 [0.50,0.78] | 0.98 [0.77,1.24] |

**Figure 7.**
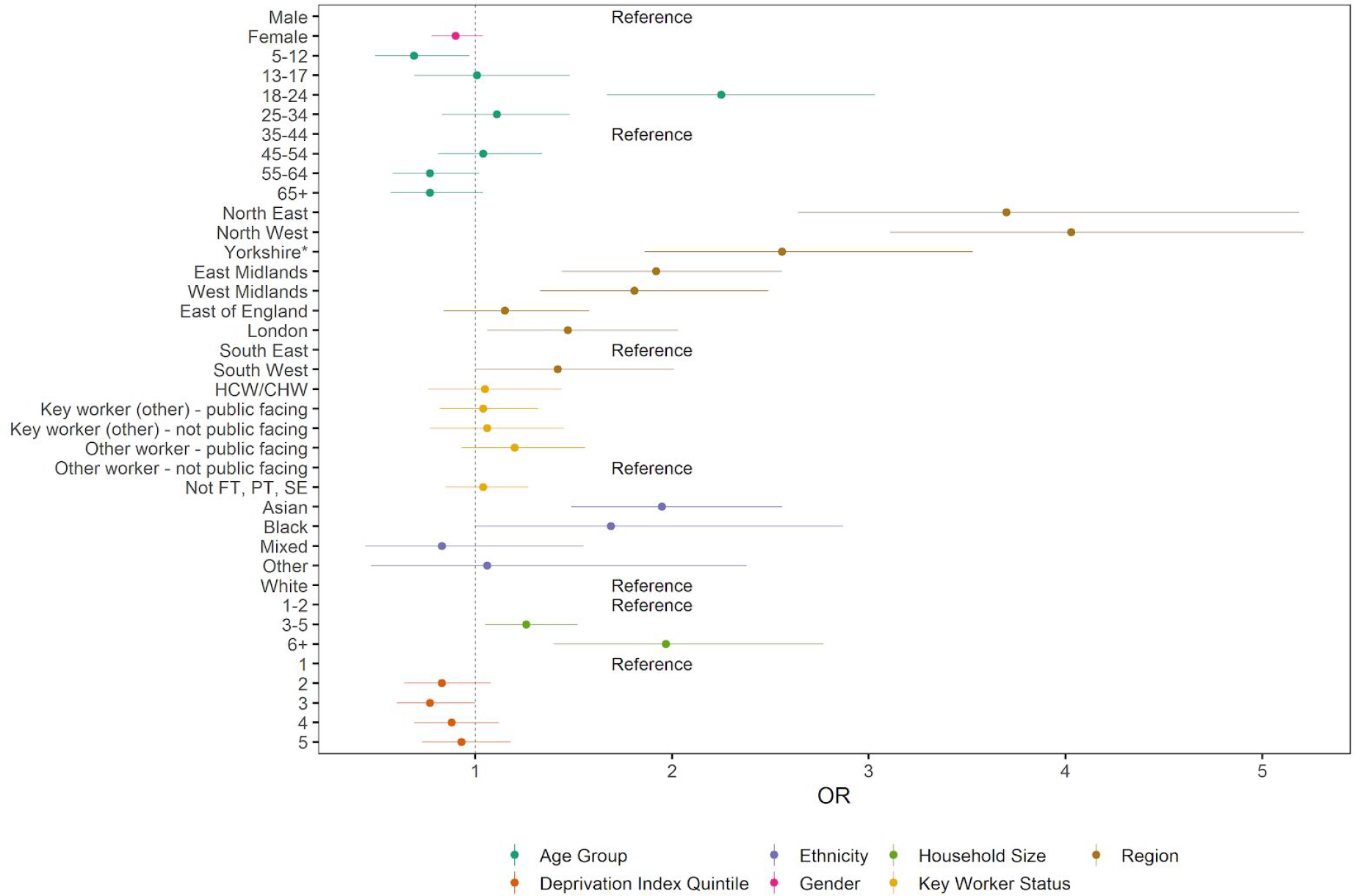
Estimated odds ratios for jointly adjusted logistic regression model of swab-positivity.

In sensitivity analyses, our point estimate of R of 1.04 (0.89, 1.21) for round 5 using data for non-symptomatic individuals was lower than the all-participant R value, although with wide confidence intervals. Other sensitivity analyses, based on more stringent criteria for positivity gave estimates of R consistent with the main findings.

## Discussion

In this fifth round of data collection from the REACT-1 study of SARS-CoV-2 virus prevalence in England, we report high and increasing prevalence of the virus especially in the North of England. At national level, around 1 in 170 people tested positive for the virus. Prevalence was highest among young adults (ages 18-24 years), but since the previous round in August / early September, it has increased markedly at all ages, including those at highest risk (ages 65 years and over). We found geographical heterogeneity of risk across England with pockets of high prevalence at LTLA level, indicating the localised as well as national nature of the pandemic.

Compared with our interim report on round 5 [5] we observed an increase in prevalence of infection in the latter part of round 5 and a firming-up of the reproduction number so that it is now reliably above one. We estimate that 450,000 (410,000, 530,000) are infected with SARS-CoV-2 in England on any one day (assuming a 75% sensitivity that a nose and throat swab picks up virus [7,8]), which translates into 45,000 new infections per day assuming virus remains detectable for 10 days on average. At least half of those with detectable virus will not display symptoms on the day of testing or previous week [5].

Our study has limitations. While we corrected for differences in population characteristics introduced by our sampling strategy and differential response rates, it is possible that our study is not fully representative of the population of England. At the time of sampling for round 5, difficulties were being experienced by symptomatic individuals in obtaining a test through the routine testing system in England. This suggests that a greater proportion of such individuals may have accessed testing through REACT-1 in round 5 than in previous rounds. Although the proportion of non-symptomatic people who tested positive was lower in the current round than previously [3], it was still 50%, and this did not vary between the first [5] and second halves of round 5 of REACT-1.

Since the rapid increase in infection that we observed at the end of August and beginning of September, a number of measures have been put into place to attempt to curb the epidemic. These include the ‘rule of six’ limiting the number of contacts at a social event to no more than six people, closure of hospitality venues by 10 pm, and a series of local lockdown measures in areas of high apparent prevalence [9]. While the rapid rate of rise observed in the previous round has slowed, the national epidemic is still growing.

In combination with the high prevalence, and evidence of faster growth in some regions, the country is now at a critical point in the second wave. Continued vigilance and adherence to the public health message of social distancing, hand washing, face covers are required, along with testing of symptomatic individuals and, if positive, isolation and rapid tracing of contacts. However, further fixed-duration measures should be considered to reduce the infection rate and limit the numbers of hospital admissions and deaths from COVID-19.

## Data Availability

The datasets generated or analysed, or both, during this study are not publicly available because of governance restrictions.

## Acknowledgements

SR, CAD acknowledge support: MRC Centre for Global Infectious Disease Analysis, National Institute for Health Research (NIHR) Health Protection Research Unit (HPRU), Wellcome Trust (200861/Z/16/Z, 200187/Z/15/Z), and Centres for Disease Control and Prevention (US, U01CK0005-01-02). GC is supported by an NIHR Professorship. PE is Director of the MRC Centre for Environment and Health (MR/L01341X/1, MR/S019669/1). PE acknowledges support from Health Data Research UK (HDR UK), the NIHR Imperial Biomedical Research Centre and the NIHR HPRUs in Environmental Exposures and Health and Chemical and Radiation Threats and Hazards, the British Heart Foundation Centre for Research Excellence at Imperial College London (RE/18/4/34215) and the UK Dementia Research Institute at Imperial (MC_PC_17114). We thank The Huo Family Foundation for their support of our work on COVID-19.

We thank key collaborators on this work -- Ipsos MORI: Kelly Beaver, Sam Clemens, Gary Welch, Nick Gilby, Andrew Cleary and Kelly Ward; Institute of Global Health Innovation at Imperial College: Gianluca Fontana, Dr Hutan Ashrafian, Sutha Satkunarajah and Lenny Naar; MRC Centre for Environment and Health, Imperial College London: Daniela Fecht; North West London Pathology and Public Health England for help in calibration of the laboratory analyses; NHS Digital for access to the NHS register; and the Department of Health and Social Care for logistic support. SR acknowledges helpful discussion with members of the UK Government Office for Science (GO-Science) Scientific Pandemic Influenza – Modelling (SPI-M) committee.

